# The Relative Importance of Mindfulness Facets and Their Interactions: Relations to Psychological Symptoms in Chronic Pain

**DOI:** 10.1101/2021.06.04.21258338

**Authors:** Zahra Azizi, Gholamreza Jandaghi, Manijeh Firoozi, Ali Zia-Tohidi, Shahrokh Ebnerasouli

## Abstract

**Objectives:** This study had three objectives: first, to investigate the relative importance of the five mindfulness facets to negative affect (NA) among patients with chronic musculoskeletal pain; second, to test the hypothesis that observing is associated with lower NA only if occurs in an accepting manner; and third, to investigate the relation between mindfulness and obsessive–compulsive symptoms (OCS).

**Methods:** One hundred and nineteen patients with chronic musculoskeletal pain filled the Five-Facets Mindfulness Questionnaire (FFMQ), the Depression, Anxiety, Stress Scale (DASS), and the Obsessive– compulsive Inventory-Revised (OCI-R). A latent variable was constructed to represent NA. Multiple regression analysis was conducted, and several indices of relative importance were calculated.

**Results:** Except for Observing, all mindfulness facets had significant bivariate and unique relation with NA. Acting with Awareness was the most important predictor, followed by Nonjudging and Describing. The contribution of Nonreactivity was small. Regarding the second objective, the Observing × Nonjudging and Observing × Nonreactivity interactions were not significant. Finally, the five facets of mindfulness explained about one-half of the variance in obsession and one-fifth of the variance in compulsion. After excluding the shared variance between obsession and compulsion, mindfulness was only related to obsession.

**Conclusions:** Except for Observing, all mindfulness facets seem to have unique contributions to psychological symptoms; among them, Acting with Awareness seems most important. Current evidence is inconsistent in supporting the moderating role of acceptance in the influence of observing. Finally, in the context of OCS, it seems that mindfulness is more related to obsession than compulsion.

---

Chronic pain (CP) is a common health condition (Breivik et al., 2006; Tsang et al., 2008), which is associated with a wide range of physical, psychological, and social consequences. It is commonly associated with anxiety and depression (Asmundson & Katz, 2009; Burke et al., 2015; Demyttenaere et al., 2007). More than half of CP patients suffer from decreased job productivity, and about one−fifth lose their job as a result of their pain condition (Breivik et al., 2006). Not surprisingly, CP is a major risk factor for suicide (World Health Organization, 2014). The destructive and disabling nature of CP along with its chronicity forces a high economic toll on societies; for instance, the annual cost of CP in the US has been estimated at $560 to $635 billion, which is far more than the total cost of heart disease or cancer (Gaskin & Richard, 2012).

The mental health conditions that are associated with CP are of major concern in its management. These are not merely sources of mental distress, but also major causes of disability (Whiteford et al., 2013) and suicide (World Health Organization, 2014). Furthermore, there is evidence that these mental health problems can exacerbate pain, or facilitate its transition from acute to chronic (Vargas-Prada & Coggon, 2015).

Numerous therapeutic approaches have been proposed for addressing psychological conditions in CP (for a review, see Kerns et al., 2011); however, based on the current evidence, their effect size is usually modest (Hoffman et al., 2007; Williams et al., 2012). The unsatisfactory effect of current therapeutic approaches has motivated the research that aims to improve the efficacy of the treatments that have shown promise. Among them, mindfulness−based interventions have earned considerable attention during the last decades, and there is now substantial evidence from large randomized trials and meta-analyses on their efficacy in improving mental health and quality of life and in reducing disability in CP patients (Cherkin et al., 2016; Hilton et al., 2016; Lauche et al., 2013; Veehof et al., 2016).

## Improving the Utility of Mindfulness

To improve the efficacy of mindfulness−based interventions, one line of research has focused on predictors of treatment outcomes, investigating that mindfulness−based treatments are (more) efficacious for whom (Gilpin et al., 2017). Another line has focused on better understanding the core processes that are responsible for the favorable effects of mindfulness. So far, the majority of the investigations on mindfulness components have been cross−sectional (Carpenter et al., 2019), and longitudinal and experimental evidence on this matter are rare (for a review and meta−analysis, see Gu et al., 2015).

The research on the differential effect of mindfulness components assumes that (a) mindfulness is a multidimensional construct, (b) some of its components may be more strongly related to some outcomes, and, (c) by putting more focus on cultivating these components, we may be able to improve the effect of mindfulness−based interventions. Furthermore, we have to assume, at least in cross−sectional investigations, that the observed relations between mindfulness facets and the outcomes of interest are due to the effect of these facets on those outcomes.

## Components of Mindfulness

In their influential article, Bishop et al. (2004) proposed a *what* and *how* framework to explain the nature of mindfulness. That is, mindfulness requires attention to the present moment (the essential *what*) and that this present−moment attention to be in a nonjudgmental and accepting manner (the essential *how*).

Baer et al. (2006) took a more empirical approach. They combined items of five well−known mindfulness measures and used this as an item pool for exploratory factor analysis. The factor analysis revealed five factors: Observing, Describing, Acting with Awareness, Nonjudging, and Nonreactivity. Then, the items were reduced to develop the Five Facet Mindfulness Questionnaire (FFMQ). This five−facet structure has been validated across numerous samples around the globe (see Karl et al., 2020, for a cross−cultural examination) and has been widely utilized for the investigation of the differential effects of mindfulness components (Carpenter et al., 2019).

## The Differential Effects of Mindfulness Components

From the development of FFMQ in 2006 (Baer et al., 2006), more than one hundred studies have used it to investigate how different mindfulness facets are related to psychological and health−related outcomes among community, student, clinical, and medical samples. Carpenter et al. (2019) conducted a comprehensive meta−analysis and synthesized the current evidence on the relation between the five facets of mindfulness and psychological symptoms, including anxiety, depression, social anxiety, and post−traumatic stress. They found that, among the five facets, Nonjudging and Acting with Awareness had the strongest relation with these symptoms (*r* = −.48 and −.47, respectively), followed by Nonreactivity and Describing (*r* = −.33 and −.29, respectively). The Observing facet, on the other hand, was almost irrelevant (*r* = .01).

One challenge in understanding the differential relation of mindfulness facets to the outcomes of interest is related to their intercorrelation. As a result of this intercorrelation, the observed bivariate effect of each facet is composed of two proportions. One proportion is its unique effect that cannot be attributed to the other facets (indicated by the partial effect size indices of the relation, when the effect of other facets has been controlled for), and the other proportion is the relationship that is shared with other facets (i.e., their bivariate correlation minus the partial relation). When the relation is shared, we cannot determine which facet is the direct contributor to the outcome, and to what extent. It is also possible that none of them be the direct contributor, rather, a common factor could be responsible for their relations. However, there are some statistical techniques, such as relative weight and dominance analysis (Nathans et al., 2012), that provide evidence about the relative importance of each predictor in the face of intercorrelation among predictors, the techniques that have rarely been utilized in this topic.

## The Theoretical Challenge in the Effect of Observing

During the previous investigations on mindfulness, an important and challenging topic has been the relationship between observing and psychological and health−related outcomes. In the initial validation of FFMQ, Baer and colleagues (2006) reported a significant positive relation between observing and worse psychological symptoms, which was unexpected, “because observing is widely described as a central feature of mindfulness” (p. 38). Similar results were found in another study on a large sample of students (Brown et al., 2015). As mentioned, in the recent comprehensive meta−analysis on more than 100 correlational studies, there was no notable relation between observing and affective symptoms (Carpenter et al., 2019). Indeed, in this meta-analysis, when student samples were only included, there was a positive significant relationship between observing and negative affective symptoms, as in Baer et al. (2006) and Brown et al. (2015).

In an attempt for explaining these unexpected results, Lindsay and Cresswell (2017) proposed the monitor and acceptance theory (MAT), which posits that the monitor and acceptance components work synergistically. The high state of awareness (i.e., monitoring), on its own, may not be associated with favorable outcomes, as it enhances all experiences, regardless of their valence. In fact, in some cases, being highly attentive may exacerbate negative feelings, which explains why during the first sessions of mindfulness training, some participants experience higher negative affect. For the monitor component to have positive effects, it needs to be in an accepting manner. Furthermore, in affective situations, the monitor process is interrupted by the ongoing emotions, unless the acceptance component works simultaneously to reduce the effect of emotion. On the other hand, the monitor component provides a scaffold for cultivating acceptance. While they noted the possibility of the independent effect of acceptance on mental and health-related outcomes, they stated that “it’s not clear how acceptance would be trained in the absence of a target object to monitor with acceptance” (p. 56).

## Current Study

The objective of this study was threefold. First, we aimed to investigate the relative importance of each mindfulness facet in the relation between mindfulness and negative affect (hereafter referred to as NA), i.e., the shared variance between different aspects of negative affective symptoms, in patients with chronic musculoskeletal pain. The shared variance between the aspects of negative affect is not only a valid construct (Goodwin, 2015; Mineka et al., 1998; Watson et al., 2008) but also, probably, a better target for research on the differential effects of mindfulness facets. There are several reasons for this proposal. First, the shared variance across different aspects of negative affective symptoms seems to constitute approximately half of their variance, and improving this shared factor may have a more widespread improvement in mental health, simultaneously affecting anxiety, depression, obsessive−compulsive symptoms, and so on. Second, current evidence suggests that mindfulness is effective across different internalizing disorders, which raises the possibility that it is influencing a common factor among these disorders. Finally, the common factor that represents the shared variance is usually associated with higher reliability, providing higher statistical power, which is especially important when studying medical or clinical populations, as the vast majority of studies on these populations do not have large samples.

An important distinction of this study is its use of relative importance analysis, which can provide a better understanding of the contribution of each mindfulness facet to psychological symptoms. While it is common to use the standardized coefficients (also called beta weight; β) to compare the importance of predictors to one another, but when the predictors are correlated (which is the case in our topic), this practice is flawed and can lead to erroneous interpretations (see Tonidandel & LeBreton, 2011). Relative importance analysis can partly overcome this shortcoming of multiple regression analysis (see Methods section).

Second, we aimed to test the suggested hypothesis that the relation between Observing and mental health is moderated by the acceptance-related facets. According to Lindsay and Cresswell (2017), among the five facets of mindfulness, as measured by FFMQ, Nonjudging and Nonreactivity are measures of acceptance-related processes.

Finally, we aimed to investigate the relation between mindfulness facets and obsessive−compulsive symptoms (OCS), as the research on this topic is rare. For instance, among the 148 studies meta-analyzed by Carpenter et al. (2019), only two had examined OCS, and none were conducted on CP patients. In general, OCS has rarely been studied among CP patients, even though some authors believe in its significance in the context of CP (Bienvenu & Cannistraro, 2002), and there is evidence on its prevalence among this population (Asmundson & Katz, 2009).

## Methods

This was a correlational study, aimed to investigate how the mindfulness facets are related to affective symptoms in patients with CP, with a focus on the relative importance of each facet. To provide a clear report, we followed the Strengthening the Reporting of Observational Studies in Epidemiology (STROBE) statement guidelines (von Elm et al., 2008).

### Setting and Participants

Participants were recruited from the pain clinic of Imam Hussein Hospital in Tehran, from September till December 2019. Adult patients (at least 18 years old), with at least a middle-school diploma, who had received a diagnosis of chronic (at least three months) musculoskeletal pain from a pain specialist were eligible. An individual was excluded if the pain was malignant or injury-related, or if the individual was under psychotropic medications or suffering from severe psychiatric conditions. Participants who met the criteria and signed a written consent were included.

### Measurement Tools

#### The Five-Facet Mindfulness Questionnaire (FFMQ)

Facets of mindfulness were assessed using the Five-Facet Mindfulness Questionnaire (FFMQ), which is a well-known 39−item measure of mindfulness (Baer et al., 2006). Several validation studies on the Iranian population have confirmed the five-factor structure of FFMQ and reported acceptable reliability and concurrent validity (e.g., Ghorbani et al., 2014; Heydarinasab et al., 2013; Sarafraz, 2016).

#### The Revised Obsessive–compulsive Inventory (OCI-R)

To assess obsessive−compulsive symptoms (OCS), the Obsessive−Compulsive Inventory-Revised (OCI-R; Foa et al., 2002) was used, a well−known 18−items scale that measures six aspects of OCS: washing, checking, ordering, obsessing, hoarding, and neutralizing. Among the Iranian population, OCI-R has shown a similar structure, with good internal consistency and test−retest reliability (Ghassemzadeh et al., 2011).

#### The Short Form Version of Depression Anxiety Stress Scales (DASS-21)

The short version of Depression Anxiety Stress Scale (Henry & Crawford, 2005) assesses the three dimensions of negative emotional states. Along with depression and anxiety, stress is assumed to be a distinct dimension of NA (Lovibond & Lovibond, 1995); however, some authors consider this subscale as a general measure of NA, not a distinct dimension (Henry & Crawford, 2005). Among the Iranian population, the DASS-21 has shown a similar structure, as well as good to excellent internal consistency and test−retest reliability (Asghari et al., 2008).

### Statistical Analysis

#### Constructing a Latent Variable for Negative Affect

We constructed a latent variable to represent the shared variance among different aspects of affective symptoms, namely, depression, anxiety, stress, and OCS. The maximum likelihood method was used for estimation. The factor scores were extracted (and multiplied by 10 to reduce the decimal places) to be the primary dependent variable in this study. This analysis was conducted in AMOS−24. The advantage of constructing such a variable is that it represents a more stable construct that underly different dimensions of common affective symptoms in CP patients, as well as it contains less measurement error, providing more power and precision for regression analysis.

#### Multiple Regression Analysis

Multiple regression analysis with marginal (Type 3) sum of squares was used to evaluate the unique contribution of each facet of mindfulness to the dependent variables. We checked all partial plots to ensure the assumptions of linearity and the constant variance have not been severely violated. Furthermore, we calculated the square root of the variance inflation factor (√VIF) to quantify the effect of collinearity on the standard error of each regression slope. We also checked studentized residuals and Mahalanobis distance values to check for univariate and multivariate outliers, respectively. If an outlier was detected, we reran the analysis leaving out the data point to see whether the results change considerably. Furthermore, we double−checked the confidence intervals of all regression slopes using the Type 4 heteroskedasticity−consistent (HC4) method, which is robust to the violation of constant variance, as well as non−normality and high leverage points (see Hayes & Cai, 2007). The regression analyses were conducted in SPSS−26.

#### Relative Importance Analysis

As mentioned in the Introduction, when the predictors are correlated, the use of the standardized coefficients to compare the importance of predictors to one another is problematic (see Tonidandel & LeBreton, 2011). To overcome this problem, we provided several indices of relative importance to facilitate a better understanding of the contribution of each mindfulness facet to NA. Commonality analysis was conducted to provide indices of the unique and common contribution of each facet to the dependent variable. Structure coefficient (*r*^2^_str_) was calculated for each predictor, which is the bivariate correlation between the predictor and the predicted values of the dependent variable. General dominance weight (GDW) was calculated, which averages the contribution of each predictor in all possible models that include the predictor. Additionally, we provided the relative weight for each predictor. RW overcomes the problem by estimating the contribution of correlated predictors by creating uncorrelated variables that are maximally related to the original variables. We presented RW in the percentile form, so it indicates the proportion of the contribution of each predictor to the total variance explained in the model; for instance, an RW of 20 for a given predictor means that 20% of the *R*^2^ can be attributed to the predictor. Finally, we conducted a series of bootstrap tests to compare the relative importance of each facet. A brief review of the common indices of relative importance is provided in Nathans et al. (2012). The indices of relative importance were calculated in R using the package “yhat” (Nimon & Oswald, 2013)

## Results

One hundred and nineteen patients with a diagnosis of chronic musculoskeletal pain participated. A summary of their demographic and pain−related characteristics is provided in Table 1. The means and *SD*s for the study variables are provided in Table 2, along with their bivariate correlations.

**Table 1.** The Summary of Participants’ Demographic and Pain−Related Characteristics

| Variable | n | % | *M* (*SD*) | Range |
| --- | --- | --- | --- | --- |
| Age |  |  | 46 (13) | 18–84 |
| 18–25 | 5 | 4 |  |  |
| 26–40 | 46 | 39 |  |  |
| 41–60 | 50 | 42 |  |  |
| 61–84 | 18 | 15 |  |  |
| Sex |  |  |  |  |
| Female | 89 | 75 |  |  |
| Male | 30 | 25 |  |  |
| Marital Status |  |  |  |  |
| Married | 99 | 83 |  |  |
| Single | 20 | 17 |  |  |
| Highest Educational Level |  |  |  |  |
| Middle–School | 42 | 35 |  |  |
| High–School | 52 | 44 |  |  |
| Tertiary | 25 | 21 |  |  |
| Pain Duration (Months) |  |  | 56 (75) | 3–480 |
| ≤ 1 Year | 45 | 38 |  |  |
| 1–3 Years | 30 | 25 |  |  |
| 3–7 Years | 19 | 16 |  |  |
| 7–15 Years | 18 | 15 |  |  |
| > 15 Years | 7 | 6 |  |  |
| Pain Intensity |  |  | 7 (2) | 2–10 |
| History of Surgery |  |  |  |  |
| Yes | 10 | 8 |  |  |
| No | 109 | 92 |  |  |

**Table 2.** Means and Standard Deviations of the Study Variables and the Bivariate Correlation Between Them

|  | *M* | *SD* | 1 | 2 | 3 | 4 | 5 | 6 | 7 | 8 | 9 | 10 | 11 |
| --- | --- | --- | --- | --- | --- | --- | --- | --- | --- | --- | --- | --- | --- |
| 1. Observing | 26.2 | 5.7 | — |  |  |  |  |  |  |  |  |  |  |
| 2. Describing | 28.8 | 6.8 | .00 | — |  |  |  |  |  |  |  |  |  |
| 3. Acting with awareness | 25.5 | 7.2 | -.03 | .36** | — |  |  |  |  |  |  |  |  |
| 4. Nonjudging | 21.5 | 6.3 | -.16 | .20* | .56** | — |  |  |  |  |  |  |  |
| 5. Nonreactivity | 23.2 | 5.8 | .23* | .20* | -.03 | -.03 | — |  |  |  |  |  |  |
| 6. OCS | 24.1 | 14.0 | .06 | -.28** | -.53** | -.48** | -.01 | — |  |  |  |  |  |
| 7. Depression | 12.1 | 10.2 | .02 | -.50** | -.50** | -.44** | -.11 | .57** | — |  |  |  |  |
| 8. Anxiety | 11.6 | 9.9 | .12 | -.38** | -.56** | -.38** | -.13 | .62** | .59** | — |  |  |  |
| 9. Stress | 17.6 | 10.9 | .10 | -.43** | -.61** | -.48** | -.23* | .68** | .72** | .74** | — |  |  |
| 10. Negative Affect<sup>(a)</sup> | 15.8 | 9.6 | .09 | -.47** | -.65** | -.52** | -.19* | .78** | .80** | .82** | .97** | — |  |
| 11. Pain Intensity | 7.3 | 2.3 | .17 | -.10 | -.15 | -.15 | .07 | .12 | .14 | .25** | .22* | .22* | — |
| 12. Pain Duration (Months) | 56.2 | 74.7 | .11 | .02 | -.06 | .13 | .00 | -.07 | -.05 | -.09 | -.04 | -.07 | .00 |

*Note.* *N* = 119. OCS = obsessive–compulsive symptoms.
<sup>a</sup>The scores for negative affect are factor scores loaded on depression, anxiety, stress, obsessions, and compulsions. The factor scores were multiplied by 10 to reduce decimals.
\**p* < .05. \*\**p* < .01.

### The Structure of the Negative Affective Symptoms

We constructed a latent variable, NA, loaded by Depression, Anxiety, Stress, and OCS. The OCS was also a latent variable loaded by its six subscales. The fit indices on this model were inconsistent, with some indicating acceptable and some indicating mediocre fit, χ^2^(26) = 62.4, *p* < .001, χ^2^ / *df* = 2.4, general fit index (GFI) = .90, comparative fit index (CFI) = .93, root mean square of error approximation (RMSEA) = .11 [.08, .14]. The inspection of the residual covariance matrix revealed a substantial error covariance between the Obsessing subscale and Depression, Anxiety, and Stress, which suggests that a substantial proportion of variance in the Obsessing subscale is not accounted for by the OCS latent factor, and this proportion of variance is correlated with the other indicators of the first−order factor, NA. We allowed Obsessing subscale to directly load on NA, leaving the OCS factor to be loaded only by compulsion subscales (e.g., washing; see the Online Resource 1 for more details on the models). The modified model showed good fit to the data, χ^2^(26) = 38, *p* = .060, χ^2^ / *df* = 1.46, GFI = .94, CFI = .98, RMSEA = .06 [.00, .10]. As our purpose was to use the factor scores of NA for further analysis, we extracted these scores from the two models to see whether there is a considerable difference between the two models. The correlation between factor scores from the two models was almost perfect, *r* = .997, *p* < .001, indicating that, for our purpose, the difference between these models is negligible; however, we used the factors scores from Model 2.

### Demographic and Pain−Related Predictors of Negative Affect

In a regression analysis, we included age, sex, marital status, history of surgery, current pain intensity, and pain duration as predictors of NA. The only significant predictor was the current pain intensity, *B* = 0.11 [0.03, 0.18], β = .25. Two multivariate outliers were detected, but excluding them did not considerably affect the results.

### The Relative Importance of Mindfulness Facets in Predicting Negative Affect

In a hierarchical linear model, we included current pain intensity in the first step and the five facets of FFMQ in the second step to predict NA. The results are presented in Table 3. In Step 1, pain intensity explained 5% of the total variance in NA. The inclusion of the five facets of mindfulness raised this value to 56%. Except for Observing, which had a nonsignificant effect, all facets had significant unique contributions to NA. Acting with Awareness had the strongest relationship, uniquely explaining 12% of the total variance in NA. Describing, Nonjudging, and Nonreactivity had fairly similar standardized slopes (−.19 to −.21). We found an influential case with a relatively high Cook’s *D*, excluding of which increased the effect of Describing from β = −.20 to β = −.27 and slightly decreased the effect of Nonjudging and Nonreactivity.

**Table 3.**
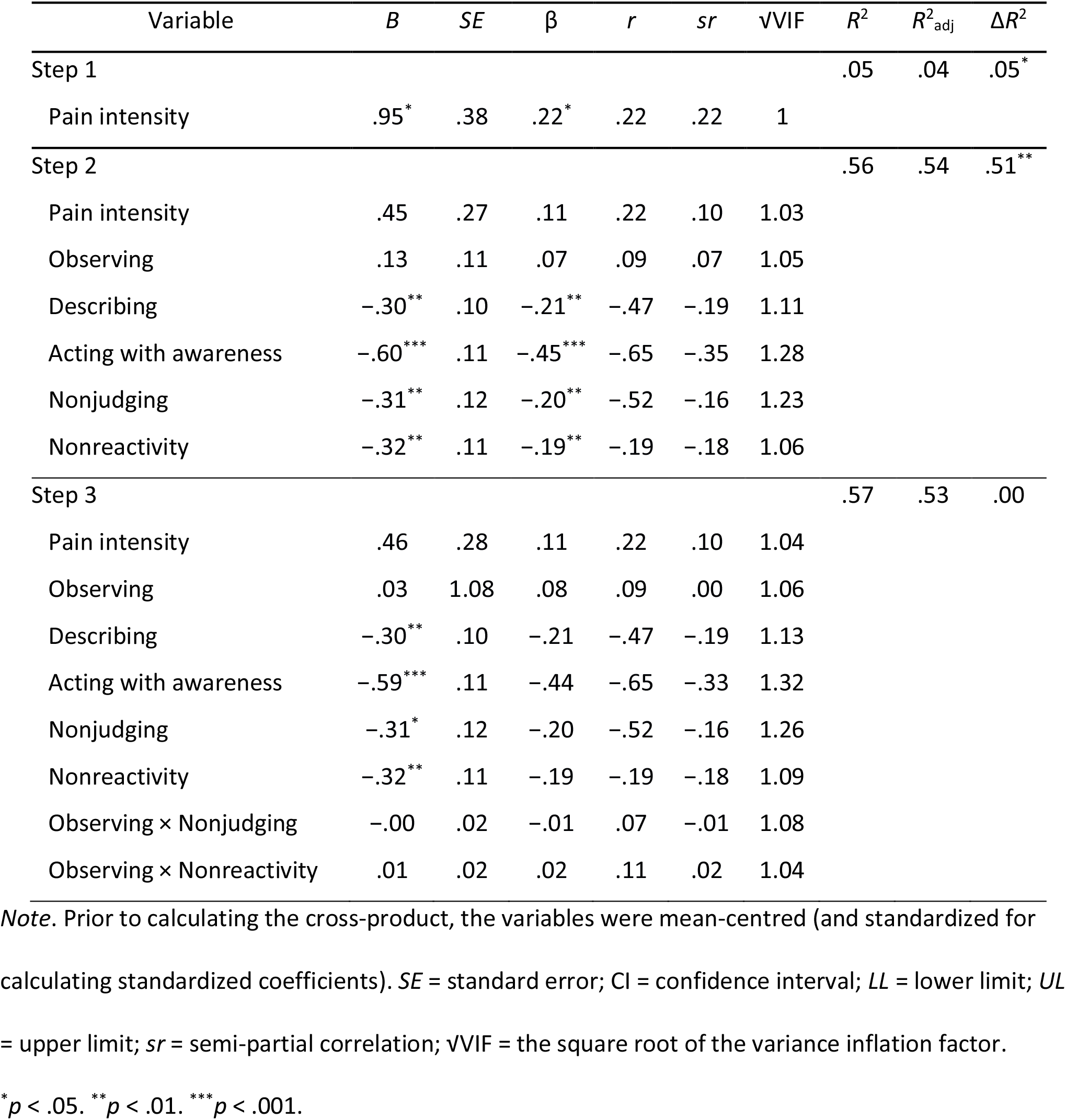
The Relationship Between the Five Facets of Mindfulness and Negative Affect

The unique and common contributions of each facet and several other indices of relative importance are provided in Table 4, and more detailed results of commonality analysis are presented in the Appendix. Acting with Awareness was the most important predictor in the model, as indicated by structure coefficients, general dominance weights (GDWs), and relative weights (RWs). It had the highest unique and common relation with NA, and its contribution was significantly higher than Observing, Nonjudging, and Nonreactivity (all bootstrap−based *p* < .05). Describing and Nonjudging had relatively similar GDW and RW. Furthermore, while the unique contribution of Nonreactivity was similar to Describing and Nonjudging, it seemed to be less important, as it did not have a notable shared contribution to NA.

**Table 4.** Relative Importance of Each Mindfulness Facet in Predicting Negative Affect

| Predictor | $\beta$ | $r^2_{\text{str}}$ | Unique | Common | GDW | RW% | Contrasts <sup>a</sup> |
| --- | --- | --- | --- | --- | --- | --- | --- |
| Observing | .06 | .01 | .00 | .00 | .00 | 1 | D, A, NJ > **O** |
| Describing | -.28 | .48 | .06 | .22 | .15 | 26 | **D** > O, NR |
| Acting with awareness | -.49 | .78 | .14 | .31 | .28 | 48 | **A** > O, NJ, NR |
| Nonjudging | -.15 | .44 | .02 | .24 | .12 | 21 | A > **NJ** > O |
| Nonreactivity | -.15 | .06 | .02 | .01 | .03 | 5 | D, A > **NR** |
| Total | – | 1.76 | .24 | .78 | .58 | 100 | – |

*Note.* $N = 118$ . $r^2_{\text{str}}$ = squared structure coefficient; GDW = general dominance weight; RW = relative weight; O = observing; D = describing; A = acting with awareness; NJ = nonjudging; NR = nonreactivity.
The results are based on a multiple regression with the five mindfulness facets as predictors of Negative Affect. One influential data point was excluded from the analysis.
<sup>a</sup> In each row, the significant contrasts to the predictor (in bold) are presented ( $p < .05$ , based on 10000 bootstrapped samples). The contrasts are between general dominance weights. Contrasting relative weights also produced similar results.

### Testing the Monitor and Acceptance Theory (MAT)

In Step 3 of the hierarchical regression model, we included Observing × Nonjudging and Observing × Nonreactivity interaction terms to test the suggested hypothesis by MAT that Observing is associated with less distress only if it occurs in an accepting manner. The change in the model fit was negligible and non-significant, Δ*R*^2^ = .001, *p* = .938 (Table 3).

To facilitate better interpretation, we conducted a simulation−based sensitivity power analysis to see for what effect size we had sufficient power (i.e., 0.8) to detect a significant interaction (The analysis was conducted in R using “SIMR” package; Green & Macleod, 2016). Based on 10000 Monte Carlo samples, we had 0.80 power to detect a standardized slope of 0.17 for Observing × Nonjudging interaction a slope of 0.18 for Observing × Nonreactivity interaction.

Furthermore, to investigate the possibility of an indirect effect of Observing on NA (although this is not suggested by MAT), we conducted a multiple mediation analysis, in which the other four facets were included as mediators. The confidence intervals for indirect effects were calculated from 10000 bootstrapped samples. The slope for the indirect effect of Observing on NA was *B* = .00 [−0.07, 0.08], β = −.00 through describing, *B* = 0.02 [−0.13, .18], β = .01 through Acting with Awareness, *B* = .06 [−0.02, 0.17], β = .03 through Nonjudging, and *B* = −.07 [−.15, −.01], β = −.04 through Nonreactivity. The total indirect effect was nonsignificant, *B* = 0.00 [−0.23, 0.27], β = .00. Considering the different directions of the indirect effects, it seems that these effects are rather random.

### The Relation Between the Aspects of Mindfulness and Obsessive−Compulsive Symptoms

Compared to general NA, depression, anxiety, and stress (see the following subsection), the relation between OCS and mindfulness facets was weaker but still substantial. As Table 5 shows, the model containing facets of mindfulness and pain intensity accounted for 34% of the total variance in OCS (*R*^2^_adj_ = 31). The effects of pain intensity, Observing and, Nonreactivity on OCS were negligible and nonsignificant, all *p* > .20. Acting with Awareness was the strongest predictor in the model, *B* = −0.67 [−1.05, −0.30], β = −.35, followed by Nonjudging, B = −0.58 [−1.00, −0.17], β = −.26. While the bivariate correlation between Describing and OCS was significant, *r* = −.28, *p* = .002, the unique contribution of Describing on OCS did not reach significance, *B =* −020 [−0.54, 0.15], β = −.10.

**Table 5.** The Relationship Between the Five Facets of Mindfulness and Obsessive–compulsive Symptoms, Depression, Anxiety, and Stress

| Variable | Obsessive–compulsive symptoms |  |  |  |  | Depression |  |  |  |  |
| --- | --- | --- | --- | --- | --- | --- | --- | --- | --- | --- |
| | *B* | *SE* | $\beta$ | *r* | *sr* | *B* | *SE* | *B* | *r* | *sr* |
| Pain intensity | .12 | .49 | .02 | .12 | .02 | .19 | .34 | .04 | .14 | .04 |
| Observing | .02 | .20 | .01 | .06 | .01 | -.04 | .14 | -.02 | .02 | -.02 |
| Describing | -.20 | .17 | -.10 | -.28 | -.09 | -.53** | .12 | -.35** | -.50 | -.32 |
| Acting with awareness | -.67** | .19 | -.35** | -.53 | -.27 | -.34* | .13 | -.24* | -.50 | -.19 |
| Nonjudging | -.58** | .21 | -.26** | -.48 | -.21 | -.39** | .14 | -.24** | -.44 | -.19 |
| Nonreactivity | -.03 | .20 | -.01 | -.01 | -.01 | -.09 | .13 | -.05 | -.11 | -.05 |
| $R^2$ | | | .34 | | | | | .42 | | |
| $R^2_{adj}$ | | | .31 | | | | | .38 | | |
| Variable | Anxiety<sup>(a)</sup> |  |  |  |  | Stress<sup>(b)</sup> |  |  |  |  |
| | *B* | *SE* | $\beta$ | *r* | *sr* | *B* | *SE* | $\beta$ | *r* | *sr* |
| Pain intensity | .67* | .33 | .15* | .25 | .15 | .51 | .33 | .11 | .22 | .10 |
| Observing | .20 | .13 | .11 | .12 | .11 | .20 | .13 | .10 | .10 | .10 |
| Describing | -.24* | .12 | -.16* | -.38 | -.15 | -.28* | .12 | -.18* | -.43 | -.16 |
| Acting with awareness | -.61** | .13 | -.44** | -.56 | -.35 | -.66** | .13 | -.44** | -.61 | -.34 |
| Nonjudging | -.09 | .14 | -.06 | -.38 | -.05 | -.29* | .14 | -.17* | -.48 | -.14 |
| Nonreactivity | -.26 | .13 | -.15 | -.13 | -.14 | -.45** | .13 | -.24** | -.23 | -.23 |
| $R^2$ | | | .40 | | | | | .51 | | |
| $R^2_{adj}$ | | | .37 | | | | | .49 | | |

Note. *SE* = standard error; *sr* = semi-partial correlation.
\* $p < .05$ . \*\* $p < .01$ . \*\*\* $p < .001$ .
<sup>a</sup> The $p$ value for nonreactivity was .054. The HC4-based $p$ value for describing was .069.
<sup>b</sup> The HC4-based $p$ value for describing and nonjudging was .062 and .074, respectively.

An important note about the OCI-R is that it is heavily weighted to measure compulsions over obsessions. To investigate whether mindfulness facets are differently related to obsessions and compulsions, we conducted two additional analyses separately for Obsession and Compulsion (i.e., the sum of the other five subscales). Interestingly, the model fit was substantially different. The model including pain intensity and mindfulness facets explained 48% of the variance in Obsession (*R*^2^_adj_ = 46), but only 24% of the variance in Compulsion (*R*^2^_adj_ = 20). As there is an overlap between obsession and compulsion, we conducted two further analyses to control for the overlapping variance. Controlling for the effect of Compulsion, mindfulness facets were still significant predictors of Obsession, *F*(5, 111) = 11.6, *p* < .001, explaining an additional 24% of its variance. On the other hand, when we controlled for the effect of Obsession, mindfulness facets were no longer significant predictors of Compulsion, *F*(5, 111) = 1.7, *p* = .136, Δ*R*^2^ = .05. These results suggest that mindfulness may be only related to obsession. However, Acting with Awareness (β = .31), Nonjudging (β = −.19), and Nonreactivity (β = −.19) were significant predictors of Obsession (all *p*s < .019).

### The Relation Between the Aspects of Mindfulness and Depression, Anxiety, and Stress

The five facets of mindfulness accounted for 41%, 38%, and 50% of the total variance in depression, anxiety, and stress, respectively. The results are presented in Table 5. Acting with Awareness and Describing facets had significant unique contributions to all three outcomes, and Observing was not significantly related to any of them.

## Discussion

The main objective of this study was to investigate how mindfulness facets are related to negative affect (NA) and obsessive−compulsive symptoms (OCS) in patients with chronic musculoskeletal pain. A total of 119 CP patients provided scores on depression, anxiety, stress, and OCS. One latent variable was constructed to represent NA, i.e., the shared variance of depression, anxiety, stress, and OCS. Except for Observing, each facet had a unique contribution in NA, altogether explaining one−half of its total variance. Acting with Awareness was the best unique predictor of NA, and the unique contributions of other three facets were of similar magnitude. Based on several relative importance indices, which take to account the unique and common contributions of predictors, Acting with Awareness was the most important predictor, followed by Describing and Nonjudging, which were also important predictors. Nonreactivity had a significant unique contribution in NA, but its overall importance was estimated to be relatively low. The relation between mindfulness facets and OCS was weaker than their relation to general NA, depression, and anxiety. However, Acting with Awareness and Nonjudging facets seemed most important to OCS, while Nonreactivity was unrelated. Furthermore, separate analyses on Obsession and Compulsion subscales showed that mindfulness may be only related to obsession. We further investigated the MAT hypothesis that the favorable effect of Observing depends on scores on acceptance−related facets, namely Nonjudging and Nonreactivity (see Lindsay & Creswell, 2017). Our data did not support this hypothesis. However, there was weak evidence on an indirect effect of Observing through Nonreactivity. In the subsequent sections, we will review the evidence on the relation of each facet to mental health outcomes among CP patients.

### The Relation Between Mindfulness Facets and Negative Affect Among CP Patients

#### The Bivariate Relations

Comparing the results from the current and previous investigations on CP patients (Jensen et al., 2018; Lee et al., 2017; Mioduszewski et al., 2018; Pleman et al., 2019; Veehof et al., 2011; Wegner et al., 2014) to the more general estimates of the bivariate relation between mindfulness facets (Carpenter et al., 2019) suggests that the relation in CP patients is fairly similar to the estimates from other medical, clinical, community, and student samples. In these studies, the bivariate correlation is usually negligible or small for Observing, small to medium for describing, medium to large for Acting with Awareness and Nonjudging, and medium for Nonreactivity. However, there are some exceptions too. In a study on a CP sample in USA, Nonreactivity had a near-zero correlation with depression (Jensen et al., 2018), which is somewhat similar to the current study. In another study on CP patients in China, in which a different version of FFMQ was used, Describing and Nonjudging had very small correlations with depression (Chen et al., 2021). However, there is some evidence that the relation between mindfulness and psychological outcomes in medical samples is stronger than community and student samples (Carpenter et al., 2019, Table 4).

#### The Unique Relations

For Describing, this and some previous studies on CP samples have supported its unique relation to lower anxiety and depression (e.g., Lee et al., 2017; Pleman et al., 2019); however, in some other studies the effect was very small and it did not reach significant (e.g., Jensen et al., 2018; Veehof et al., 2011). For Acting with Awareness and Nonjudging, the findings seem to consistently support their unique contributions to psychological symptoms (e.g., Jensen et al., 2018; Lee et al., 2017; Pleman et al., 2019; Veehof et al., 2011). In this study, and several previous studies, Acting with Awareness had the strongest unique contribution to psychological outcomes (e.g., Pleman et al., 2019; Veehof et al., 2011). For Nonreactivity, the evidence is inconsistent, with some studies reporting significant unique relation (e.g., Pleman et al., 2019; Veehof et al., 2011) and some not (Jensen et al., 2018; Lee et al., 2017); in this study, Nonreactivity had significant unique contributions to general NA and stress. Notably, the bivariate and the unique relation between Nonreactivity and psychological outcomes were similar, as Nonreactivity was fairly unrelated to the other influential facets of mindfulness.

#### The Relative Importance

In this study, we supplemented the multiple regression analysis with a series of relative importance analyses, which combine the unique and the shared contribution of each facet to estimate its relative importance in the model. Among the four influential facets of mindfulness (i.e., excluding Observing), Acting with Awareness seemed to be the most important aspect of mindfulness in its relation with NA, while Nonreactivity seemed the least important. To our knowledge, none of the previous studies on CP patients has conducted relative importance analysis, and this type of analysis among other populations is also rare. In one study on a US community sample, Acting with Awareness, Describing, and Nonjudging were more important than Nonreactivity in predicting the general wellbeing of individuals without regular practice of meditation; while for whom with regular practice, Nonreactivity was more important than Nonjudging (Hanley et al., 2015).

### The Effect of Observing and the Moderating Role of Acceptance

So far, at least eight investigations have tested the hypothesis that the favorable effects of monitoring depend on the level of acceptance (Lindsay & Creswell, 2017). A summary of these investigations is presented in Table 6. The evidence is mixed. However, the pattern of results may suggest two points. First, the measurement seems to play a role. While four out of the seven investigations on Observing × Nonreactivity interaction have yielded significant results, none of the four investigations on Observing × Nonjudging interaction reach significance (all are subscales of FFMQ). It seems that the issue of measuring acceptance needs further investigation. While both Nonjudging and Nonreactivity have been considered as measures of acceptance-related processes (Lindsay & Creswell, 2017), in this and some previous studies, the correlations between these two facets were near-zero or even negative (e.g., Brown et al., 2015; Hou et al., 2014), which raises doubt whether they share an underlying common factor (i.e., acceptance). Second, culture may also play a role; four out of the five studies that support this hypothesis have been conducted in the US, while three studies conducted in Italy, Japan, and Iran (current study) have not supported this hypothesis.

**Table 6.**
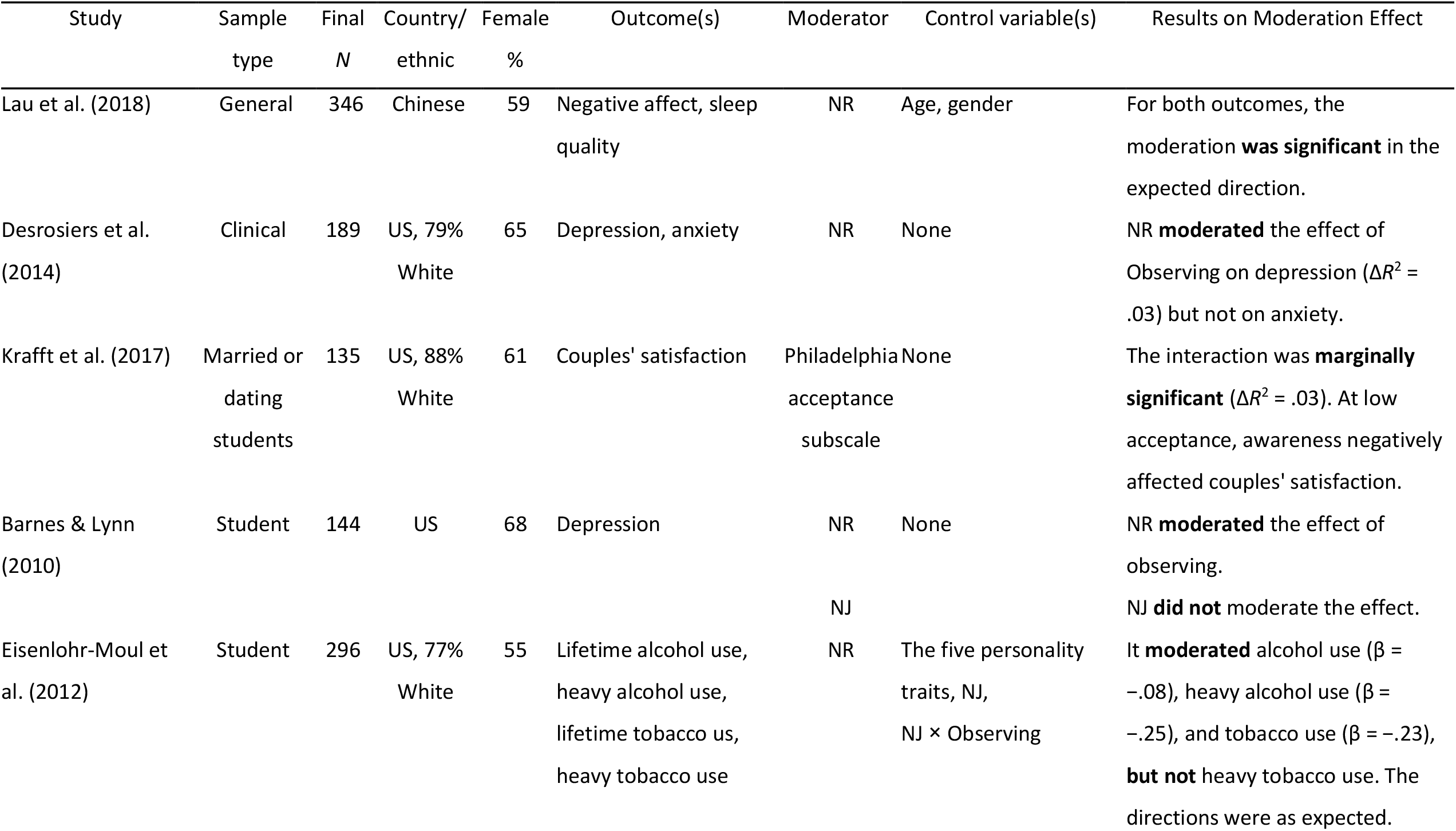
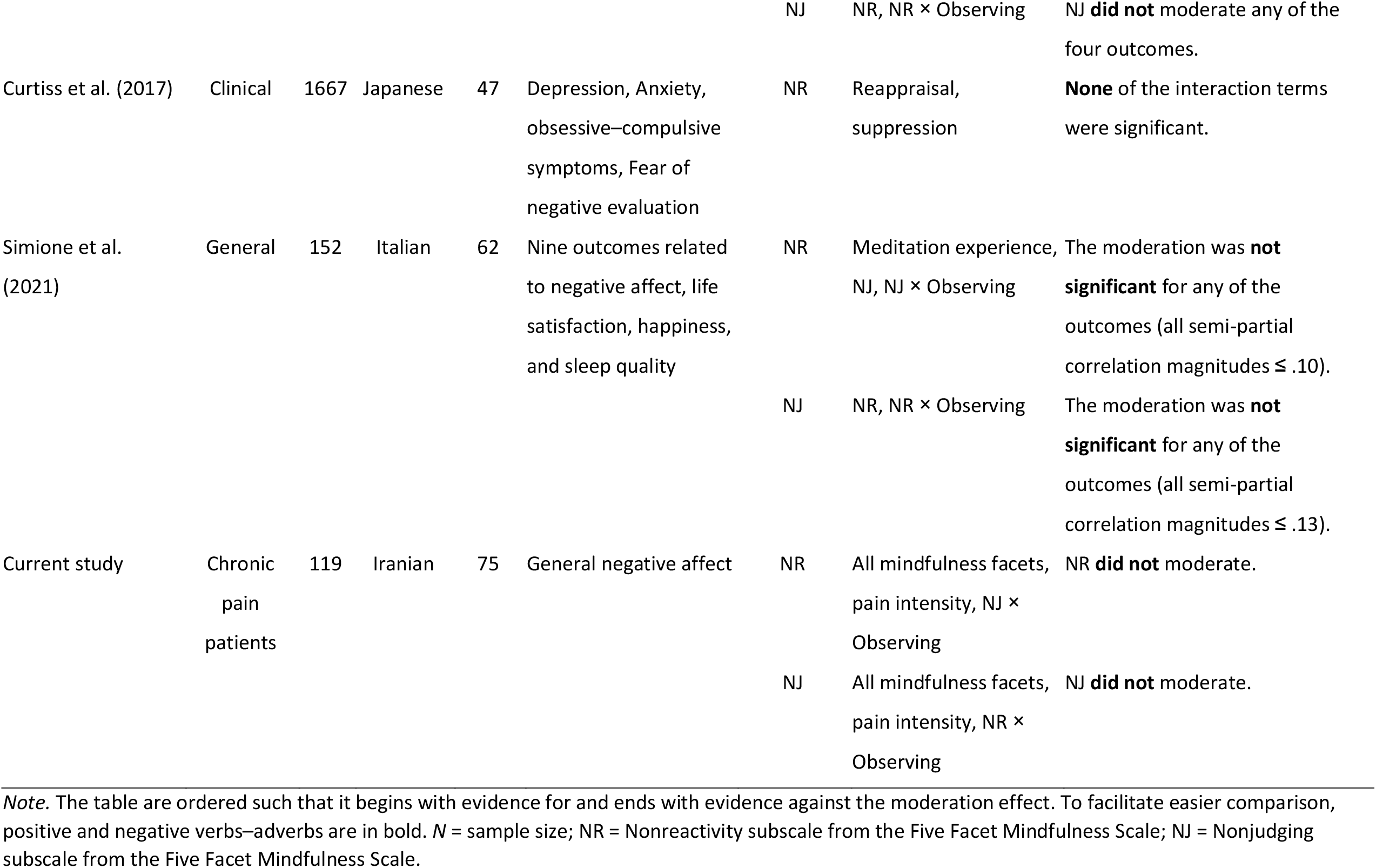
Previous Findings on the Moderation Effect of Acceptance on Monitoring

Besides the interaction hypothesis which is suggested to explain the unexpected relation between observing and psychological symptoms, another attempt has focused on the measurement of the monitor component. Rudkin and colleagues (2018) hypothesized that the Observing facet of FFMQ does not sufficiently capture the observing construct. They conducted an exploratory factor analysis on a pool of items related to this component, which revealed three correlated subfactors: Body Observing, Emotion Awareness, and External Perception. Among these three factors, only Emotion Awareness was notably related to worry, stress, and anxiety. They suggested that the absence of items that assess emotional awareness may be the reason that the FFMQ Observing facet has shown unexpected relation to mental health.

### The Differential Relation of Mindfulness to Obsession and Compulsion

While the investigation on the relation between mindfulness and mental health has mostly focused on depression and anxiety, during the last 5 years several investigations have documented the relation between mindfulness and OCS, even though none have been conducted on CP patients. Five correlational studies reported significant small to medium negative bivariate correlations between the mindfulness facets and OCS, except for Observing (Curtiss et al., 2017; Emerson et al., 2018; Leeuwerik et al., 2020; Solem et al., 2015). In two comparative studies, patients with OCD scored lower on the mindfulness facets than healthy controls, except for Observing (Didonna et al., 2019) or Observing and Nonreactivity (Crowe & McKay, 2016). In the current study, OCS had negative small to medium correlations with mindfulness facets, except for Observing and Nonreactivity.

An important finding in this study was the difference between obsession and compulsion in how they related to mindfulness. Mindfulness accounted for one−half of the variance in obsession and about one−fifth of the variance in compulsion. Furthermore, when the shared variance between obsession and compulsion was controlled for, mindfulness still was an important predictor of obsession, but not of compulsion. Supporting this argument, in an investigation on a large clinical sample (*N* = 1871), mindfulness had a large correlation with the obsession subscale of OCI-R (*r* = −.52), but its relations to other subscales were small (ranging from −.21 to −.27; Leeuwerik et al., 2020, Appendix 3), and given the reported medium to large correlation coefficients between obsession and compulsion subscales, a substantial proportion of the observed relation between mindfulness and compulsion subscales can be attributed to the shared variance between compulsion subscales and obsession. In another large sample study (*N* = 583), mindfulness had a significant medium−sized correlation with the frequency of intrusive thoughts (*r* = .40), but its relation with behavioral compulsions (i.e., washing, checking, ordering, and repeating) was small (*r* = −.16, Emerson et al., 2018). These results raise the possibility that, while patients with OCD usually score significantly lower on mindfulness scales (Didonna et al., 2019), this construct may only be related to their mental obsession.

The differential relation of mindfulness with obsession and compulsion may have an implication for the research on this topic. Some OCS measures, such as OCI-R, put excessive weight on compulsion over obsession (Foa et al., 2002), and this can underestimate the relation between OCS and mindfulness.

## Limitations

It is important to note that the observed relation between mindfulness facets can, to some extent, be functions of confounding variables. For instance, large correlations have been reported between Describing and emotional intelligence (*r* = .60) and between Nonjudging and neuroticism (*r* = −.55; Baer et al., 2006). In a recent investigation on the incremental effect of mindfulness, after controlling for the effects of the Big−Five personality traits, mindfulness facets showed small effects (Tran et al., 2020). Emotion regulation strategies are other important confounding variables (e.g., see Curtiss et al., 2017). Although it may be practically impossible to include all important confounding variables in a single investigation, one might be aware of these research limitations when interpreting the results.

## Supporting information

Online Resource 1

## Data Availability

None.

## Declarations

### Ethical Approval

The ethical approval for this study was obtained from the University of Shahid Beheshti (IR.SBMU.RETECH.REC.1398.331).

### Informed Consent

All participants provided a written informed consent.

## Appendix

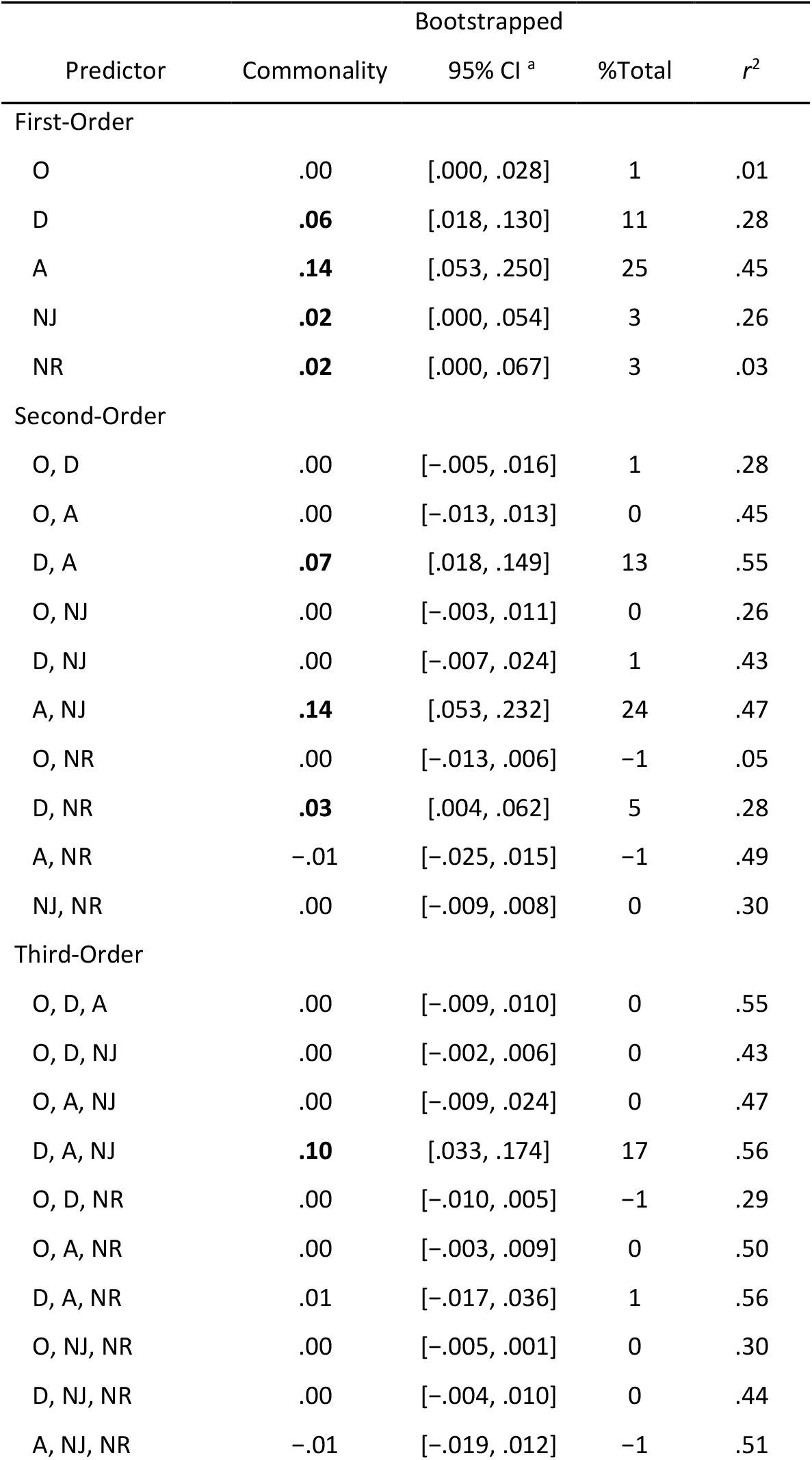
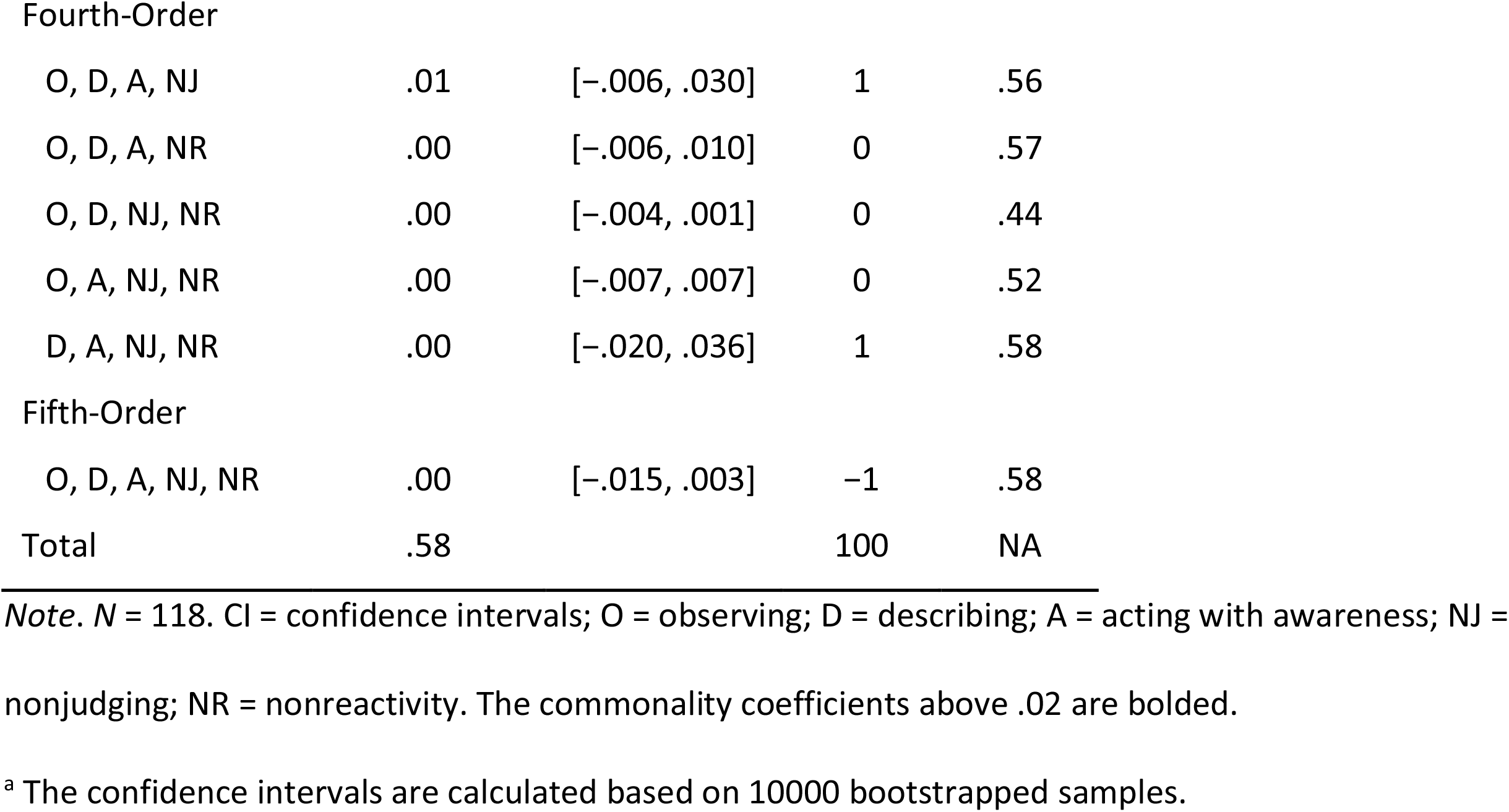
The Detailed Results of Commonality Analysis of the Relation Between Mindfulness Facets and Negative Affect

| Predictor | Commonality | Bootstrapped |  |  |
| --- | --- | --- | --- | --- |
|  |  | 95% CI <sup>a</sup> | %Total | *r*<sup>2</sup> |
| First-Order |  |  |  |  |
| O | .00 | [.000, .028] | 1 | .01 |
| D | **.06** | [.018, .130] | 11 | .28 |
| A | **.14** | [.053, .250] | 25 | .45 |
| NJ | **.02** | [.000, .054] | 3 | .26 |
| NR | **.02** | [.000, .067] | 3 | .03 |
| Second-Order |  |  |  |  |
| O, D | .00 | [−.005, .016] | 1 | .28 |
| O, A | .00 | [−.013, .013] | 0 | .45 |
| D, A | **.07** | [.018, .149] | 13 | .55 |
| O, NJ | .00 | [−.003, .011] | 0 | .26 |
| D, NJ | .00 | [−.007, .024] | 1 | .43 |
| A, NJ | **.14** | [.053, .232] | 24 | .47 |
| O, NR | .00 | [−.013, .006] | −1 | .05 |
| D, NR | **.03** | [.004, .062] | 5 | .28 |
| A, NR | −.01 | [−.025, .015] | −1 | .49 |
| NJ, NR | .00 | [−.009, .008] | 0 | .30 |
| Third-Order |  |  |  |  |
| O, D, A | .00 | [−.009, .010] | 0 | .55 |
| O, D, NJ | .00 | [−.002, .006] | 0 | .43 |
| O, A, NJ | .00 | [−.009, .024] | 0 | .47 |
| D, A, NJ | **.10** | [.033, .174] | 17 | .56 |
| O, D, NR | .00 | [−.010, .005] | −1 | .29 |
| O, A, NR | .00 | [−.003, .009] | 0 | .50 |
| D, A, NR | .01 | [−.017, .036] | 1 | .56 |
| O, NJ, NR | .00 | [−.005, .001] | 0 | .30 |
| D, NJ, NR | .00 | [−.004, .010] | 0 | .44 |
| A, NJ, NR | −.01 | [−.019, .012] | −1 | .51 |
| Fourth-Order |  |  |  |  |
| O, D, A, NJ | .01 | [−.006, .030] | 1 | .56 |
| O, D, A, NR | .00 | [−.006, .010] | 0 | .57 |
| O, D, NJ, NR | .00 | [−.004, .001] | 0 | .44 |
| O, A, NJ, NR | .00 | [−.007, .007] | 0 | .52 |
| D, A, NJ, NR | .00 | [−.020, .036] | 1 | .58 |
| Fifth-Order |  |  |  |  |
| O, D, A, NJ, NR | .00 | [−.015, .003] | −1 | .58 |
| Total | .58 |  | 100 | NA |

*Note.* $N = 118$ . CI = confidence intervals; O = observing; D = describing; A = acting with awareness; NJ = nonjudging; NR = nonreactivity. The commonality coefficients above .02 are bolded.
<sup>a</sup> The confidence intervals are calculated based on 10000 bootstrapped samples.

