## Supplementary material for "The Relative Importance of Mindfulness Facets and Their Interactions: Relations to Psychological Symptoms in Chronic Pain": Online Resource 1

**Online Resource 2**

*The Structure of the Primary Dependent Variable*

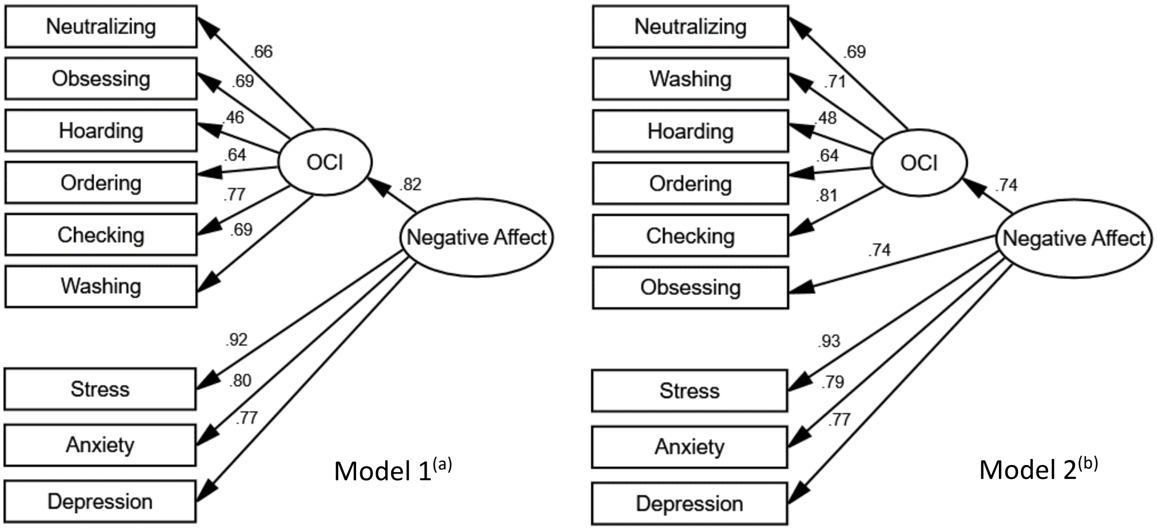


*Note*. N = 119. The maximum likelihood method was used for estimation. While the models differed in model fit, the correlation of factor scores for the Negative Affect from the two models was almost perfect, $r = .997$, $p < .001$.

[a] $\chi^2(26) = 62$, $p < .001$, CMIN/DF = 2.40, GFI = .90, CFI = .93, RMSEA = .11 (90% CI: .08, .14).

[b] $\chi^2(26) = 38$, p = .060, CMIN/DF = 1.46, GFI = .94, CFI = .98, RMSEA = .06 (90% CI: .00, .10).
